# Distinct Sex-Associated Phenotypes of Sudden Cardiac Arrest Across Clinical, ECG, and Echocardiographic Domains: Replication in Two Communities and Comparison with the Framingham Heart Study

**DOI:** 10.64898/2026.10.30.26366963

**Authors:** Thien Tan Tri Tai Truyen, Harpriya Chugh, David Nguyen, Kotoka Nakamura, Audrey Uy-Evanado, Emelia J. Benjamin, Kyndaron Reinier, Sumeet S. Chugh

## Abstract

**Background:** Sex differences in sudden cardiac arrest (SCA) incidence and outcomes are well established, but risk stratification underperforms in women. It is important to delineate sex-specific patterns in SCA across clinical, electrocardiographic, and echocardiographic domains.

**Methods:** We studied out-of-hospital SCA cases from the Oregon Sudden Unexpected Death Study (ORSUDS; Portland, 2002–2020; discovery) and Prediction of Sudden Death in Multi-Ethnic Communities (PRESTO; Ventura County, 2015–2024; external replication). Sex differences in 38 comorbidities, 19 ECG and 12 echocardiographic measures were estimated using multivariable-adjusted logistic regression. Female sex was the modeled dependent measure (male as reference). ORSUDS estimates were compared with those in the Framingham Heart Study (FHS), a community-based reference cohort, using FDR-adjusted Cochran Q tests.

**Results:** Among 5130 ORSUDS cases, 1715 (33.4%) were women (median age, 70.4 years [IQR, 58.7–81.8]) and 3415 (66.6%) were men (65.6 [55.0–76.8]). Compared with men, women had higher BMI (per 5 kg/m², 1.11 [1.08–1.15]) and higher odds of thyroid disease (2.56 [2.17–3.13]), asthma (2.33 [1.92–2.86]), mood disorder (2.17 [1.85–2.50]), dementia (1.27 [1.02–1.56]), and left ventricular hypertrophy (LVH, 3.13 [2.44–4.00]). Women also had lower odds of coronary artery disease (0.61 [0.52–0.71]), atrial fibrillation (0.67 [0.56–0.79]), prolonged QTc (0.45 [0.35–0.57]), prolonged PR (0.53 [0.41–0.68]), premature contractions (0.77 [0.60–0.98]), sleep apnea (0.53 [0.43–0.65]), and reduced LVEF (0.40 [0.29–0.54]). Most findings were replicated in PRESTO (n=2719). In contrast, several of these differences were reversed in the community (FHS, n=3539;FDR-significant): prolonged QTc was more common in women, whereas LVH, dementia, heart failure, and higher BMI were more common in men.

**Conclusions:** Sex differences among SCA cases were mostly reproducible in SCA cohorts, and distinct from sex-related patterns in the community. These results have implications for development of sex-specific SCA risk assessment and prevention strategies.

**Clinical Perspective:** *What is new?:*

- This study is the first community-based analysis to comprehensively characterize pre-arrest sex-associated profiles across clinical, ECG, and echocardiographic domains, externally validate them, and compare them with a community reference population.
- Women with SCA were more strongly associated with non-cardiac comorbidities, LVH, preserved LVEF, and mitral valve abnormalities, whereas male cases were characterized by coronary and peripheral vascular disease, atrial fibrillation, prolonged PR and QTc intervals, premature contractions, and reduced LVEF.
- Several sex associations, including QTc prolongation, LVH, body mass index, heart failure, and dementia, differed in magnitude or direction from patterns in the community reference population.

*What are the clinical implications?:*

- Our findings suggest candidate features for future sex specific risk factor analyses and potential components for sex-specific risk models for SCA.
- Clinicians may need to recognize the critical need to extend beyond dependence on just the value of the left ventricular ejection fraction for SCA risk prediction, especially in women.

## Introduction

Sudden cardiac arrest (SCA) remains a major contributor to cardiovascular mortality despite advances in prevention, emergency response, and post-arrest care. Sex differences in epidemiology of SCA have been consistently reported across community-based and nationwide cohorts. Men experience substantially higher SCA incidence than women across most adult age groups, although this difference narrows with advancing age.^1–4^ These differences suggest that sex is an important determinant of SCA risk, and women who have SCA may have clinical characteristics that differ from those observed in men.^5^

Women are generally older at the time of arrest, less likely to have documented coronary artery disease (CAD) or severe left ventricular systolic dysfunction, and are more likely to present with non-shockable rhythms, unwitnessed events, and arrest at home.^6–10^ Autopsy-based studies have also reported that women have less obstructive CAD and a higher proportion of nonischemic myocardial disease, myocardial fibrosis, or non-cardiac causes.^11,12^ However, other cohort studies have identified conventional cardiovascular risk factors, including hypertension, diabetes, smoking, obesity or central adiposity, prior myocardial infarction (MI), heart failure (HF), left ventricular hypertrophy (LVH), and QT-related abnormalities, as important predictors of sudden cardiac death (SCD) in women.^13–15^ These findings indicate that traditional cardiovascular risk factors remain relevant in women, but their distribution and clinical expression may differ from those in men.

Therefore, comprehensive studies of sex differences in SCA are needed with evaluation of detailed clinical comorbidity profiles, prearrest electrocardiographic features, and echocardiographic findings. Such analyses should be externally validated to determine whether sex differences among SCA cases are specific to SCA or whether they simply reflect background sex differences present in the community population.

To address this knowledge gap, we evaluated sex-associated SCA profiles before the index event, across three domains: clinical comorbidities, electrocardiographic measures, and echocardiographic findings. We used the Oregon Sudden Unexpected Death Study (ORSUDS) as the discovery cohort and the Prediction of Sudden Death in Multi-Ethnic Communities (PRESTO) study as an external replication cohort, with the Framingham Heart Study (FHS) serving as a community-cohort reference. We hypothesized that SCA cases exhibit reproducible sex-associated profiles across clinical, electrical, and structural modalities, and that selected sex differences among SCA cases diverge from sex patterns observed in a community cohort.

## Methods

### Study design and population selection of discovery cohort

For discovery, we utilized the ORSUDS study, which involved 5854 SCA cases occurring between 2002 and 2020 in the Portland, Oregon metro area. This is an ongoing community-based study encompassing a catchment population of about 1 million. The methodology, detailed elsewhere,^6,16,17^ involves identifying SCA cases through emergency medical services (EMS), including ambulance and fire department first responders. For each potential SCA case, medical history was retrieved from medical records, EMS narratives, and autopsy findings when available. A panel of three physicians adjudicated the SCA cases using the following definition: SCA was a sudden, pulseless collapse due to a likely cardiac etiology, occurring rapidly after symptom onset when witnessed or if unwitnessed, within 24 h of the individual being last seen in their usual state of health.^18^ Non-cardiac causes—such as trauma, drowning, overdose, terminal illness, active malignancies, or extracardiac causes like pulmonary embolism and other natural causes—were systematically excluded.

### Variables definition and data preprocessing

To comprehensively evaluate sex differences among SCA cases, we employed three complementary analytic approaches: (1) demographic and comorbidity characteristics; (2) ECG measures among SCA cases with a pre-event ECG; and (3) echocardiographic findings among SCA cases with a pre-event echocardiogram. The specific variables included in each approach are detailed in the Supplementary methods. Variables with more than 20% missingness were excluded.

### Study design and population selection for external replication cohort and community cohort referent

To assess the generalizability and robustness of our findings, we performed an external replication using data from the ongoing PRESTO study from February 1, 2015, to January 31, 2024, which covers a catchment population of ∼850,000 in Ventura County, CA. In this study, SCA cases were identified using methods similar to those employed in the ORSUDS study, including consistent approaches to case ascertainment, adjudication, inclusion criteria, data retrieval, and SCA definitions. The methodology of the PRESTO study has been described in previously published studies. For the validation population, we selected SCA cases aged 18 years or older with comprehensive medical records, including ECG and echocardiography results obtained prior to SCA.

As the community cohort referent, we used the Offspring cohort of the Framingham Heart Study (FHS), a prospective, multigenerational epidemiological study designed to identify factors that contribute to cardiovascular disease. The study methodology and participant selection criteria have been described previously.^19–21^ The Offspring cohort, started in 1971, comprised 5124 individuals, including the children of the Original Cohort and the spouses of those children. Follow-up examinations occurred every 4 to 8 years.^22^ For the present analysis, we included participants who survived to Exam 7 (1998–2001), providing a contemporary reference cohort that temporally aligns with the initiation of ORSUDS.

### Statistical analysis

In this case-case analysis, initial univariable analyses were conducted without imputation. Continuous variables were presented as mean ± standard deviation (SD) and, where appropriate, median with interquartile range (IQR), while categorical variables were reported as counts and percentages. Group comparisons employed the chi-square test for proportions and either Student’s t test or the Mann–Whitney U test for continuous variables, as appropriate. To obtain adjusted ORs accounting for potential confounders, we performed multivariable logistic regression with female sex as the dependent measure (vs male sex), with multiple imputations via chained equations (MICE; R package *mice*) to address missing data. Predictive mean matching was used to impute continuous variables, and logistic regression was applied to impute binary variables; the number of imputed datasets (m) was set to 10 to enhance robustness. All variables within each analytic approach were included in the multivariable models, which were fit across all 10 imputed datasets; the numbers of covariates were 38 for demographic and clinical, 19 for ECG, and 12 for echo. Final adjusted ORs and their 95% confidence intervals (CIs) were derived by pooling results across the imputed datasets to reduce uncertainty and bias.

For each feature, we estimated the female-versus-male log OR and its standard error separately in ORSUDS, PRESTO, and FHS. We assessed heterogeneity in the sex association across the three cohorts using Cochran’s *Q*test. The inverse-variance-weighted common-effect estimate, calculated using weights (1/SE²), served only as the reference under the null hypothesis of a shared association and was not interpreted as a pooled effect.^23,24^ Two pairwise comparisons were performed: ORSUDS vs. PRESTO (validation) and ORSUDS vs. FHS (community cohort reference). A non-significant Q (p > 0.05) indicated that the two estimates were statistically compatible — the sex difference was consistent across cohorts — whereas a significant Q (p < 0.05) indicated heterogeneity in the magnitude or direction of the sex effect. To account for multiple testing across features, Q p-values were adjusted using the Benjamini–Hochberg false discovery rate (FDR) procedure.^25^ Specifically, for the ORSUDS vs. PRESTO comparison, a non-significant Q supported reproducibility of the sex difference in the validation cohort; for the ORSUDS-vs-FHS comparison, an FDR-adjusted p < 0.05 identified features whose sex association in SCA cases diverged from the community cohort pattern. All analyses were performed using SPSS and R, with a two-sided p-value < 0.05 considered statistically significant.

## Results

### Sex differences in demographic and clinical characteristics of SCA cases

In the ORSUDS study, we included 5130 out of 5854 total SCA cases with medical records and demographic information available for clinical and demographic analyses, which comprised 1715 women with SCA (33.4%) and 3415 men with SCA (66.6%) (Figure 1). In the multivariable-adjusted model, advanced age, higher mean body mass index (BMI), diabetes, chronic kidney disease (CKD), thyroid disease, autoimmune disease, asthma, chronic pain, dementia, mood disorder, and calcium channel blocker use were associated with female sex. In contrast, White race, heavy alcohol use, illicit drug use, CAD, atrial fibrillation (AF), hyperlipidemia, peripheral vascular disease, sleep apnea, and anemia were associated with male sex. Liver disease showed a borderline association with male sex (Table 1).

**Figure 1.**
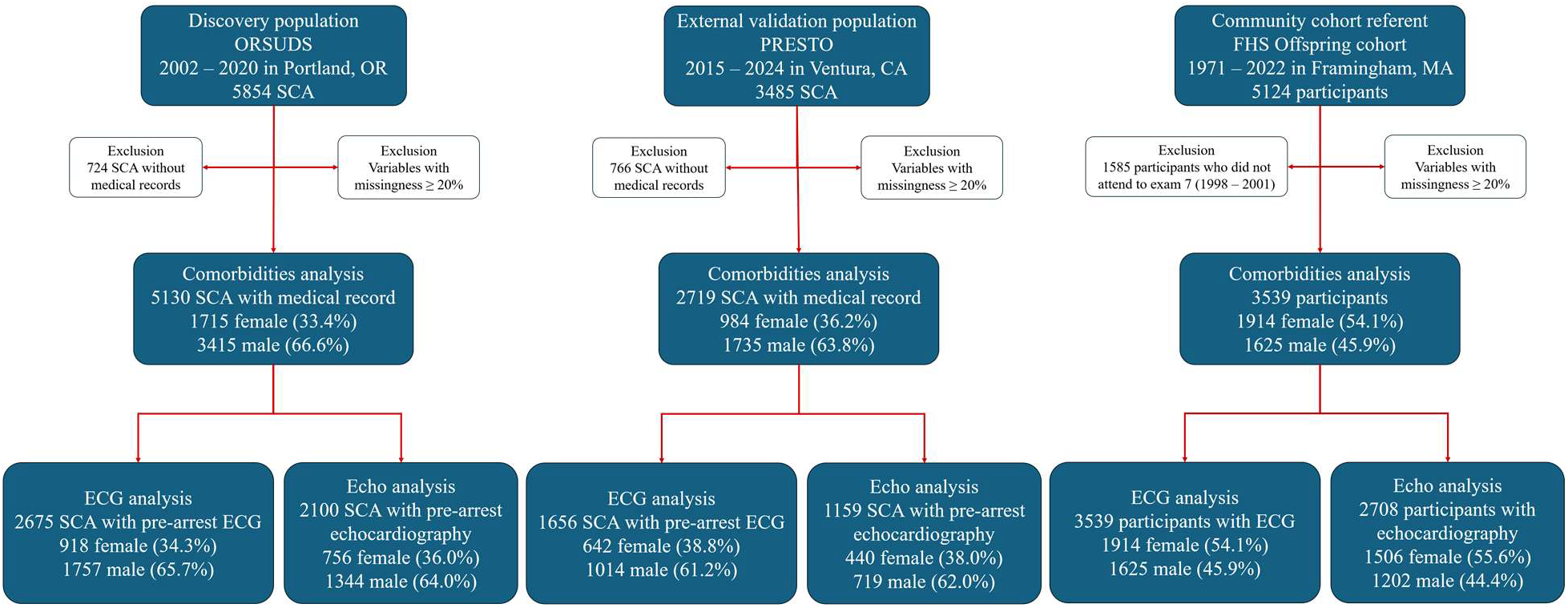
Flowchart of sudden cardiac arrest cases selection in discovery and validation population. The discovery cohort included sudden cardiac arrest (SCA) cases identified through the Oregon Sudden Unexpected Death Study (ORSUDS) in the Portland, Oregon, metropolitan area from 2002 through 2020. Of 5854 identified cases, 724 were excluded because medical records were unavailable, resulting in 5130 cases for final analysis. Variables with at least 20% missing data were also excluded. Three analytic samples were defined: The comorbidity analysis included all 5,130 cases with available medical records; The ECG analysis included the subset with a pre- arrest ECG (n = 2675); and the echocardiographic analysis included the subset with a pre-arrest echocardiogram (n = 2100). For external replication, we used SCA cases identified through the Prediction of Sudden Death in Multi-Ethnic Communities (PRESTO) study in Ventura County, California, from 2015 through 2024. Of 3485 identified cases, 2719 met the criteria for the comorbidity analysis and had available medical records. The ECG and echocardiographic analyses included the subsets with a pre-arrest ECG (n = 1656) and a pre-arrest echocardiogram (n = 1159), respectively. As the community cohort referent, we used the Offspring cohort of the Framingham Heart Study (FHS) in Massachusetts, from 1971 to 2022. For the present analysis, we included participants who survived to Exam 7 (1998–2001), providing a contemporary reference cohort that temporally aligns with the initiation of ORSUDS. In FHS, the clinical comorbidity and ECG comparisons included 3539 FHS participants comprising 1914 women (54.1%) and 1625 men (45.9%), whereas the echocardiographic analysis included 2708 participants, comprising 1506 women (55.6%) and 1202 men (44.4%). The same eligibility criteria and variable-level missingness threshold were applied in all cohorts.

**Table 1.**
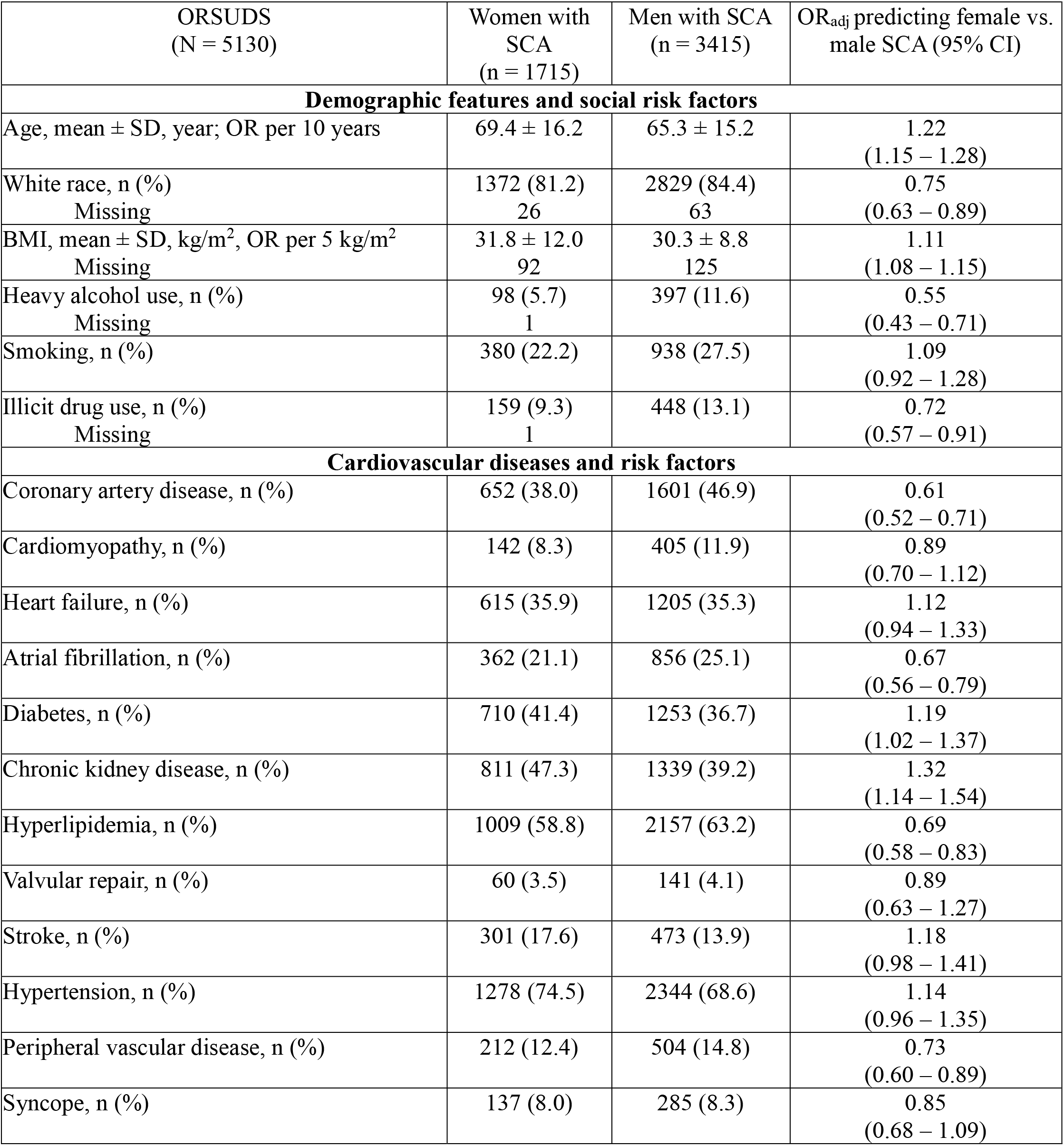
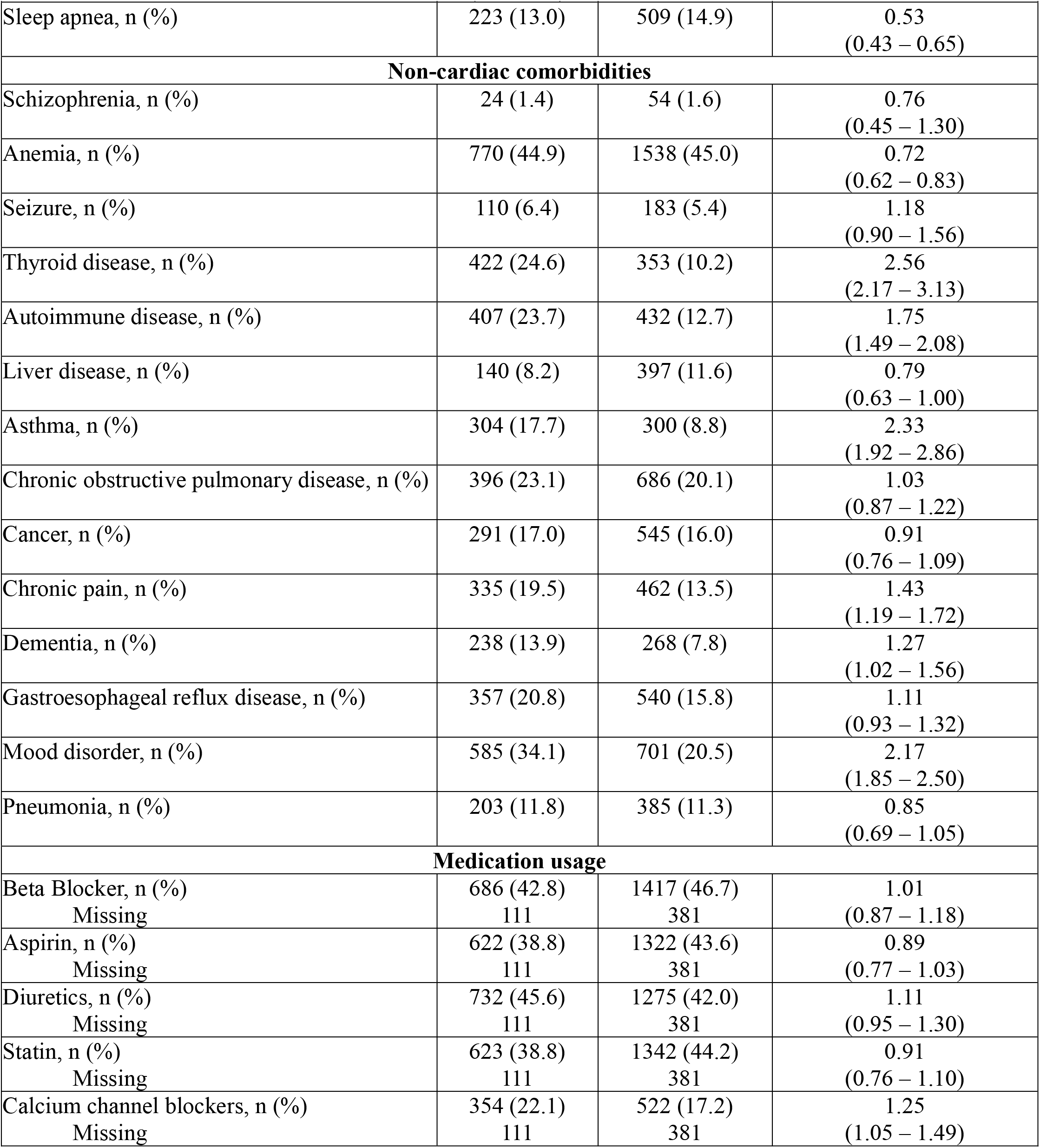
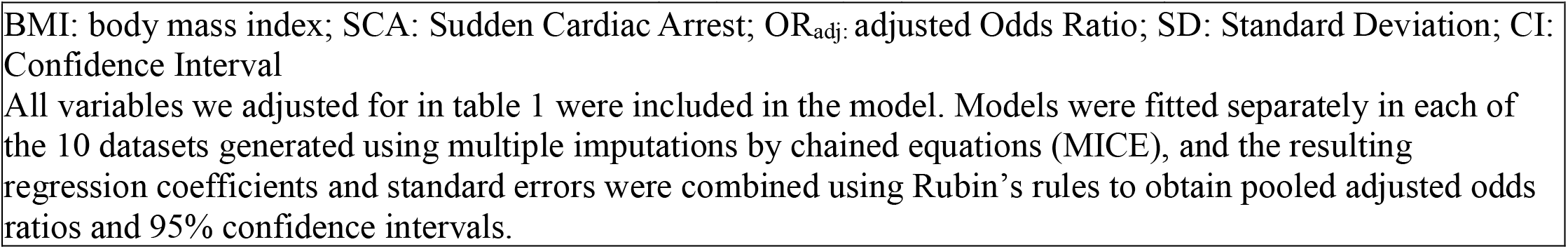
Demographic and clinical characteristics of sudden cardiac arrest cases across sexes.

The ECG analysis included 2675 SCA cases with a pre-arrest ECG, comprising 918 female cases (34.3%) and 1757 male cases (65.7%). After multivariable adjustment, higher P-wave axis values, longer corrected JT (JTc), ECG-defined LVH, prolonged Tpeak–Tend (Tpe), and delayed QRS transition were associated with female sex. Bradycardia, prolonged PR, prolonged corrected QT (QTc), longer T-wave duration, and premature contractions were associated with male sex. (Table 2).

**Table 2.** Sex differences in electrocardiogram measures in sudden cardiac arrest cases.

| ORSUDS<br>(N = 2675) | Women with SCA<br>(n = 918) | Men with SCA<br>(n = 1757) | OR<sub>adj</sub> predicting female vs. male SCA (95% CI) |
|---|---|---|---|
| Age, mean ± SD, year, OR per 10 years | 70.6 ± 15.1 | 68.0 ± 14.0 | 1.32<br>(1.22 – 1.41) |
| White race, n (%)<br>Missing | 710 (78.1)<br>9 | 1441 (82.9)<br>18 | 0.83<br>(0.66 – 1.03) |
| BMI, mean ± SD, kg/m<sup>2</sup>, OR per 5 kg/m<sup>2</sup><br>Missing | 32.3 ± 12.6<br>19 | 30.5 ± 8.9<br>30 | 1.16<br>(1.12 – 1.22) |
| Normal heart rate, n (%)<br>Bradycardia, n (%)<br>Tachycardia, n (%)<br>Missing | 704 (76.8)<br>97 (10.6)<br>116 (12.6)<br>1 | 1291 (73.5)<br>252 (14.3)<br>214 (12.2) | Reference<br>0.60 (0.45 – 0.79)<br>1.00 (0.76 – 1.32) |
| Prolonged PR (≥200 ms), n (%)<br>Missing | 125 (14.6)<br>64 | 350 (22.0)<br>169 | 0.53<br>(0.41 – 0.68) |
| Intraventricular conduction delay, n (%) | 228 (24.8) | 564 (32.1) | 0.89<br>(0.71 – 1.14) |
| Axis P wave, median (IQR), OR per 10 degrees<br>Missing | 55 (38 – 67)<br>112 | 51 (33 – 66)<br>253 | 1.05<br>(1.01 – 1.09) |
| Left ventricular hypertrophy, n (%) | 244 (26.6) | 219 (12.5) | 3.13<br>(2.44 – 4.00) |
| Delayed intrinsic deflection (≥50 ms), n (%)<br>Missing | 193 (24.9)<br>142 | 465 (30.5)<br>234 | 0.85<br>(0.67 – 1.06) |
| Prolonged Tpeak-Tend, n (%)<br>Missing | 501 (56.5)<br>32 | 850 (49.7)<br>46 | 1.41<br>(1.12 – 1.79) |
| T wave duration, mean ± SD, ms, OR per 10 ms<br>Missing | 185 ± 39<br>116 | 183 ± 41<br>193 | 0.96<br>(0.93 – 0.99) |
| Corrected JT interval, mean ± SD, ms, OR per 10 ms<br>Missing | 359 ± 40<br>1 | 346 ± 41<br>6 | 1.14<br>(1.11 – 1.18) |
| Prolonged corrected QT, n (%)<br>Missing | 540 (58.9)<br>1 | 1067 (60.9)<br>6 | 0.45<br>(0.35 – 0.57) |
| QRS-T angle, median (IQR), OR per 10 degrees<br>Missing | 52 (21–113)<br>14 | 56 (22–120)<br>26 | 0.98<br>(0.96 – 1.00) |
| Delayed QRS transitional, n (%)<br>Missing | 385 (43.2)<br>27 | 619 (36.1)<br>42 | 1.25<br>(1.03 – 1.54) |
| Premature contraction, n (%)<br>Missing | 133 (14.6)<br>7 | 302 (17.3)<br>13 | 0.77<br>(0.60 – 0.98) |
| QRS axis abnormalities, n (%)<br>Missing | 202 (22.1)<br>5 | 436 (24.9)<br>9 | 0.96<br>(0.76 – 1.22) |
| Atrial enlargement, n (%)<br>Missing | 228 (26.8)<br>66 | 381 (24.1)<br>179 | 1.06<br>(0.86 – 1.30) |

Body mass index: body mass index; ms: milliseconds; SCA: Sudden cardiac arrest; OR<sub>adj:</sub> adjusted Odds Ratio; SD: Standard Deviation; CI: Confidence Interval
All variables we adjusted for in table 2 were included in the model. Models were fitted separately in each of the 10 datasets generated using multiple imputations by chained equations (MICE), and the resulting regression coefficients and standard errors were combined using Rubin’s rules to obtain pooled adjusted odds ratios and 95% confidence intervals.

**Table 3.** Sex differences in echocardiographic measures in sudden cardiac arrest cases.

| ORSUDS<br>(N = 2100) | Women with SCA<br>(n = 756) | Men with SCA<br>(n = 1344) | OR<sub>adj</sub> predicting female vs. male SCA (95% CI) |
|---|---|---|---|
| Age, mean ± SD, year, OR per 10 years | 71.3 ± 14.3 | 69.1 ± 13.6 | 1.19<br>(1.11 – 1.28) |
| White race, n (%)<br>Missing | 574 (76.6)<br>7 | 1102 (83.5)<br>25 | 0.64<br>(0.50 – 0.81) |
| BMI, mean ± SD, kg/m<sup>2</sup>, OR per 5 kg/m<sup>2</sup><br>Missing | 32.7 ± 13.0<br>8 | 31.0 ± 9.3<br>12 | 1.15<br>(1.10 – 1.20) |
| LAd, mean ± SD, mm<br>Missing | 40.2 ± 6.6<br>151 | 44.3 ± 7.5<br>239 | NA |
| LAE, n (%)<br>Missing | 206 (34.2)<br>154 | 330 (30.0)<br>244 | 1.47<br>(1.14 – 1.92) |
| LVEDd, mean ± SD, mm<br>Missing | 46.4 ± 8.9<br>103 | 52.5 ± 9.9<br>204 | NA |
| LV enlarged, n (%)<br>Missing | 88 (13.5)<br>106 | 182 (16.0)<br>209 | 1.05<br>(0.75 – 1.49) |
| IVSd, mean ± SD, mm<br>Missing | 11.4 ± 2.6<br>101 | 11.9 ± 2.6<br>202 | NA |
| LVPWd, mean ± SD, mm<br>Missing | 11.2 ± 2.4<br>101 | 11.5 ± 2.3<br>202 | NA |
| LV mass, mean ± SD, g<br>Missing | 196.6 ± 74.4<br>103 | 251.3 ± 91.1<br>204 | NA |
| LVH, n (%)<br>Missing | 239 (36.8)<br>103 | 338 (29.8)<br>202 | 1.79<br>(1.41 – 2.22) |
| LVEF, mean ± SD, %<br>Missing | 55.1 ± 15.2<br>4 | 48.4 ± 16.3<br>9 | NA |
| LVEF Classification, n (%)<br>Preserved (≥ 50%)<br>Mid-range (36-49%)<br>Reduced (≤35%)<br>Missing | 540 (71.8)<br>112 (14.9)<br>100 (13.3)<br>4 | 729 (54.6)<br>306 (22.9)<br>300 (22.5)<br>9 | Reference<br>0.49 (0.37 – 0.65)<br>0.40 (0.29 – 0.54) |
| SWMA, n (%)<br>Missing | 175 (26.7)<br>101 | 473 (41.4)<br>202 | 0.67<br>(0.52 – 0.85) |
| Mitral abnormalities, n (%) | 116 (15.3) | 164 (12.2) | 1.39<br>(1.05 – 1.85) |
| Aortic abnormalities, n (%) | 78 (10.3) | 121 (9.0) | 0.88<br>(0.63 – 1.20) |

SCA: Sudden Cardiac Arrest; OR<sub>adj:</sub> adjusted Odds Ratio; SD: Standard Deviation; CI: Confidence Interval; BMI: body mass index; LAd: Left Atrium Diameter; LAE: Left Atrial Enlargement; LVEDd: Left Ventricular End Diastolic Diameter; LV: Left Ventricular; IVSd: Intraventricular Septum Diameter; LVPWd: Left Ventricular Posterior Wall Diameter; LVH: Left Ventricular Hypertrophy; LVEF: Left Ventricular Ejection Fraction; SWMA: Segmental Wall Motion Abnormalities; NA: Not available.
All variables we adjusted for in table 3 were included in the model. Models were fitted separately in each of the 10 datasets generated using multiple imputations by chained equations (MICE), and the resulting regression coefficients and standard errors were combined using Rubin’s rules to obtain pooled adjusted odds ratios and 95% confidence intervals.

The echocardiographic analysis included 2100 SCA cases with pre-arrest echocardiography, comprising 756 female cases (36.0%) and 1344 male cases (64.0%). After multivariable adjustment, left atrial enlargement, LVH, and mitral valve abnormalities were associated with female sex. Compared with preserved left ventricular ejection fraction (LVEF), both mid-range and reduced LVEF were associated with male sex as well as segmental wall-motion abnormalities.

### External replication using PRESTO study cohort

The PRESTO external replication cohort included 2719 SCA cases, comprising 984 female cases (36.2%) and 1735 male cases (63.8%). Baseline characteristics and adjusted results for the three analytic approaches are presented in Tables S1–S3. Overall, the sex-associated patterns identified in ORSUDS were broadly reproduced in PRESTO. Across three approaches, FDR-adjusted Cochran Q tests showed no evidence of between-cohort heterogeneity for 36 of 38 clinical variables, 18 of 19 ECG variables, and all 12 variables included in the echocardiographic comparison (Table 4).

**Table 4.** FDR-adjusted Cochran Q tests to show heterogeneity of adjusted Odds Ratios for women with SCA (vs. men with SCA) between ORSUDS and PRESTO population.

| Features | ORSUDS adjusted OR (95% CI) | PRESTO adjusted OR (95% CI) | FDR adjusted Cochran’s Q test p value |
|---|---|---|---|
| **Clinical characteristics** |  |  |  |
| Age per 10 years | 1.22 (1.15 – 1.28) | 1.16 (1.08 – 1.25) | 0.71 |
| BMI per 5 kg/m<sup>2</sup> | 1.11 (1.08 – 1.15) | 1.03 (0.97 – 1.09) | 0.20 |
| White race | 0.75 (0.63 – 0.89) | 0.85 (0.65 – 1.14) | 0.79 |
| Heavy alcohol use | 0.55 (0.43 - 0.71) | 0.59 (0.40 - 0.89) | 0.92 |
| Smoking | 1.09 (0.92 - 1.28) | 1.01 (0.75 - 1.35) | 0.90 |
| Illicit drug use | 0.72 (0.57 - 0.91) | 0.73 (0.51 - 1.05) | 0.94 |
| Coronary artery disease | 0.61 (0.52 - 0.71) | 0.58 (0.48 - 0.71) | 0.91 |
| Cardiomyopathy | 0.89 (0.70 - 1.12) | 0.83 (0.62 - 1.12) | 0.91 |
| Heart failure | 1.12 (0.94 - 1.33) | 1.33 (1.06 - 1.69) | 0.71 |
| Atrial fibrillation | 0.67 (0.56 - 0.79) | 0.65 (0.52 - 0.81) | 0.94 |
| Diabetes | 1.19 (1.02 - 1.37) | 0.85 (0.70 - 1.03) | 0.09 |
| Chronic kidney disease | 1.32 (1.14 - 1.54) | 0.88 (0.71 - 1.09) | 0.05 |
| Hyperlipidemia | 0.69 (0.58 - 0.83) | 0.83 (0.64 - 1.09) | 0.71 |
| Valvular repair | 0.89 (0.63 - 1.27) | 0.76 (0.50 - 1.16) | 0.86 |
| Stroke | 1.18 (0.98 - 1.41) | 1.01 (0.78 - 1.28) | 0.71 |
| Hypertension | 1.14 (0.96 - 1.35) | 1.12 (0.88 - 1.43) | 0.94 |
| Peripheral vascular disease | 0.73 (0.60 - 0.89) | 0.88 (0.66 - 1.19) | 0.71 |
| Syncope | 0.85 (0.68 - 1.09) | 0.70 (0.51 - 0.96) | 0.74 |
| Sleep apnea | 0.53 (0.43 - 0.65) | 0.47 (0.33 - 0.65) | 0.86 |
| Schizophrenia | 0.76 (0.45 - 1.30) | 1.11 (0.38 - 3.23) | 0.86 |
| Anemia | 0.72 (0.62 - 0.83) | 1.41 (1.12 - 1.75) | <0.001 |
| Seizure | 1.18 (0.90 - 1.56) | 1.61 (1.10 - 2.33) | 0.71 |
| Thyroid disease | 2.56 (2.17 - 3.13) | 2.27 (1.85 - 2.86) | 0.76 |
| Autoimmune disease | 1.75 (1.49 - 2.08) | 2.33 (1.72 - 3.13) | 0.59 |
| Liver disease | 0.79 (0.63 - 1.00) | 0.48 (0.32 - 0.71) | 0.20 |
| Asthma | 2.33 (1.92 - 2.86) | 1.52 (1.12 - 2.04) | 0.17 |
| Chronic obstructive pulmonary disease | 1.03 (0.87 - 1.22) | 1.20 (0.95 - 1.52) | 0.71 |
| Cancer | 0.91 (0.76 - 1.09) | 0.95 (0.76 - 1.20) | 0.91 |
| Chronic pain | 1.43 (1.19 - 1.72) | 1.18 (0.95 - 1.47) | 0.71 |
| Dementia | 1.27 (1.01 - 1.56) | 1.47 (1.12 - 1.92) | 0.76 |
| Gastroesophageal reflux disease | 1.11 (0.93 - 1.32) | 1.03 (0.81 - 1.32) | 0.90 |
| Mood disorder | 2.17 (1.85 - 2.50) | 2.38 (1.89 - 3.03) | 0.84 |
| Pneumonia | 0.85 (0.69 - 1.04) | 0.88 (0.68 - 1.16) | 0.92 |
| Beta Blocker | 1.01 (0.87 - 1.18) | 1.30 (1.06 - 1.59) | 0.24 |
| Aspirin | 0.89 (0.77 - 1.03) | 0.88 (0.72 - 1.08) | 0.94 |
| Diuretics | 1.11 (0.95 - 1.28) | 1.09 (0.88 - 1.33) | 0.94 |
| Statin | 0.91 (0.76 - 1.09) | 0.83 (0.65 - 1.06) | 0.86 |
| Calcium channel blockers | 1.25 (1.05 - 1.49) | 1.33 (1.08 - 1.64) | 0.90 |
| **ECG features** |  |  |  |
| Age per 10 years | 1.32 (1.22 - 1.41) | 1.35 (1.23 - 1.47) | 0.86 |
| White race | 0.83 (0.66 - 1.03) | 0.96 (0.78 - 1.20) | 0.78 |
| Body mass index per 5 kg/m<sup>2</sup> | 1.16 (1.11 - 1.22) | 1.09 (1.02 - 1.16) | 0.49 |
| Normal heart rate<br>Bradycardia<br>Tachycardia | Reference<br>0.60 (0.45 - 0.79)<br>1.00 (0.76 - 1.32) | Reference<br>0.79 (0.55 - 1.16)<br>1.04 (0.76 - 1.41) | 0.73<br>0.86 |
| Prolonged PR interval | 0.53 (0.41 - 0.68) | 0.49 (0.35 - 0.68) | 0.86 |
| Intraventricular conduction delay | 0.89 (0.71 - 1.14) | 0.62 (0.47 - 0.83) | 0.49 |
| Axis P wave per 10 degrees | 1.04 (1.01 - 1.09) | 1.01 (0.98 - 1.05) | 0.65 |
| Left ventricular hypertrophy | 3.03 (2.44 - 3.85) | 3.70 (2.63 - 5.26) | 0.78 |
| Delayed intrinsic deflection | 0.85 (0.67 - 1.06) | 0.81 (0.61 - 1.08) | 0.86 |
| Prolonged Tpeak-Tend | 1.41 (1.12 - 1.79) | 0.80 (0.61 - 1.05) | 0.04 |
| T wave duration per 10 ms | 0.96 (0.93 - 0.99) | 0.96 (0.93 - 1.00) | 0.86 |
| Corrected JT interval per 10 ms | 1.14 (1.11 - 1.18) | 1.12 (1.09 - 1.16) | 0.86 |
| Prolonged corrected QT interval | 0.45 (0.35 - 0.57) | 0.46 (0.34 - 0.62) | 0.86 |
| QRS-T angle per 10 degrees | 0.98 (0.96 - 1.00) | 1.01 (0.98 - 1.03) | 0.53 |
| Delayed QRS transitional zone | 1.25 (1.03 - 1.54) | 1.10 (0.87 - 1.41) | 0.78 |
| Premature contraction | 0.77 (0.60 - 0.98) | 0.61 (0.44 - 0.85) | 0.73 |
| QRS axis abnormalities | 0.96 (0.75 - 1.22) | 0.90 (0.69 - 1.18) | 0.86 |
| Atrial enlargement | 1.06 (0.86 - 1.30) | 1.12 (0.83 - 1.54) | 0.86 |
| **Echocardiographic findings** |  |  |  |
| Age per 10 years | 1.19 (1.11 - 1.28) | 1.14 (1.03 - 1.27) | 0.61 |
| White race | 0.64 (0.50 - 0.81) | 0.96 (0.75 - 1.23) | 0.10 |
| BMI per 5 kg/m<sup>2</sup> | 1.15 (1.10 - 1.20) | 1.02 (0.94 - 1.11) | 0.10 |
| Left ventricular enlargement | 1.05 (0.75 - 1.49) | 0.63 (0.40 - 0.97) | 0.22 |
| Left ventricular hypertrophy | 1.79 (1.41 - 2.22) | 2.00 (1.52 - 2.70) | 0.61 |
| Ejection Fraction Classification<br>Preserved (≥ 50%)<br>Mid-range (36-49%)<br>Reduced (≤35%) | Reference<br>0.49 (0.37 - 0.65)<br>0.40 (0.29 - 0.54) | Reference<br>0.69 (0.48 - 0.99)<br>0.45 (0.29 - 0.70) | 0.37<br>0.62 |
| Segmental Wall Motion Abnormality | 0.67 (0.52 - 0.85) | 0.81 (0.56 - 1.18) | 0.61 |
| Mitral abnormalities, n (%) | 1.39 (1.05 - 1.85) | 1.69 (1.19 - 2.38) | 0.61 |
| Aortic abnormalities, n (%) | 0.88 (0.63 - 1.20) | 0.74 (0.48 - 1.14) | 0.61 |

BMI: Body mass index

Specifically, the overall phenotype was preserved, and women with SCA were characterized by a non-cardiac comorbidity profile and men with SCA predominantly by a cardiovascular and behavioral risk profile (Table 4). Significant between-cohort heterogeneity after FDR correction was limited to anemia and CKD. Anemia reversed from male association in ORSUDS to female association in PRESTO (OR = 0.72 versus 1.41; Cochran’s Q = 24.47; FDR-adjusted heterogeneity p < 0.001). CKD was associated with female sex in ORSUDS, whereas the point estimate suggested an association with male sex in PRESTO (OR = 1.32 versus 0.88; Q = 9.20; FDR-adjusted heterogeneity p = 0.05). Diabetes also showed associations in opposite directions across cohorts, but the corresponding heterogeneity did not remain significant after FDR correction (OR = 1.19 versus 0.85; FDR-adjusted heterogeneity p = 0.09).

The ECG analysis included 1656 SCA cases, comprising 642 female cases (38.8%) and 1014 male cases (61.2%). The principal electrical phenotype was reproduced: ECG-defined LVH and corrected JT intervals remained more common among female cases, whereas male cases continued to have a higher prevalence of prolonged PR, QTc intervals, and premature contractions (Table 4). A prolonged Tpeak-to-Tend interval was the only ECG feature showing significant between-cohort heterogeneity after FDR correction. It was associated with female sex in ORSUDS, whereas the point estimate suggested an association with male sex in PRESTO (OR = 1.41 versus 0.80; Cochran’s Q = 9.40; FDR-adjusted heterogeneity p = 0.04).

The echocardiographic analysis included 1159 SCA cases, comprising 440 female cases (38.0%) and 719 male cases (62.0%). The structural phenotype observed in ORSUDS was also reproduced in PRESTO: LVH and mitral valve abnormalities were more common among female cases, whereas mildly reduced and reduced LVEF were more common among male cases (Table 4). None of the 12 variables in the echocardiographic models showed significant between-cohort heterogeneity after FDR correction (all FDR-adjusted heterogeneity p ≥ 0.05).

### Comparison of sex differences in SCA vs. community cohort using FHS

In FHS, the clinical comorbidity and ECG comparisons included 3539 FHS participants comprising 1914 women (54.1%) and 1625 men (45.9%), whereas the echocardiographic analysis included 2708 participants, comprising 1506 women (55.6%) and 1202 men (44.4%). The characteristics for each approach have been described in supplemental materials (Supplemental Results and Tables S4-S6).

FDR-adjusted Cochran Q tests identified significant ORSUDS–FHS heterogeneity for 14 of the 25 clinical and medication features (Figure 2, Table S7). For eight features, the ORSUDS and FHS point estimates were opposite, including age, BMI, HF, hypertension, valvular disease or repair, dementia, pneumonia, and calcium channel blocker use. Notably, older age and higher BMI were associated with female sex in the ORSUDS clinical model but with male sex in FHS (age: Q=29.40, FDR-adjusted p <0.001; BMI: Q=55.56, FDR-adjusted p <0.001. HF changed from a nonsignificant female-directed estimate in ORSUDS to a significant male association in FHS (OR 1.12 versus 0.41; Q=5.95, FDR-adjusted p = 0.03), while dementia changed from female association in ORSUDS to male association in FHS (OR 1.27 versus 0.16; Q=8.85, FDR- adjusted p = 0.007). Significant heterogeneity for AF, CKD, asthma, thyroid disease, aspirin use, and diuretic use reflected differences in estimated effect magnitude rather than direction. The remaining 11 clinical features showed no evidence of significant between-cohort heterogeneity.

**Figure 2.**
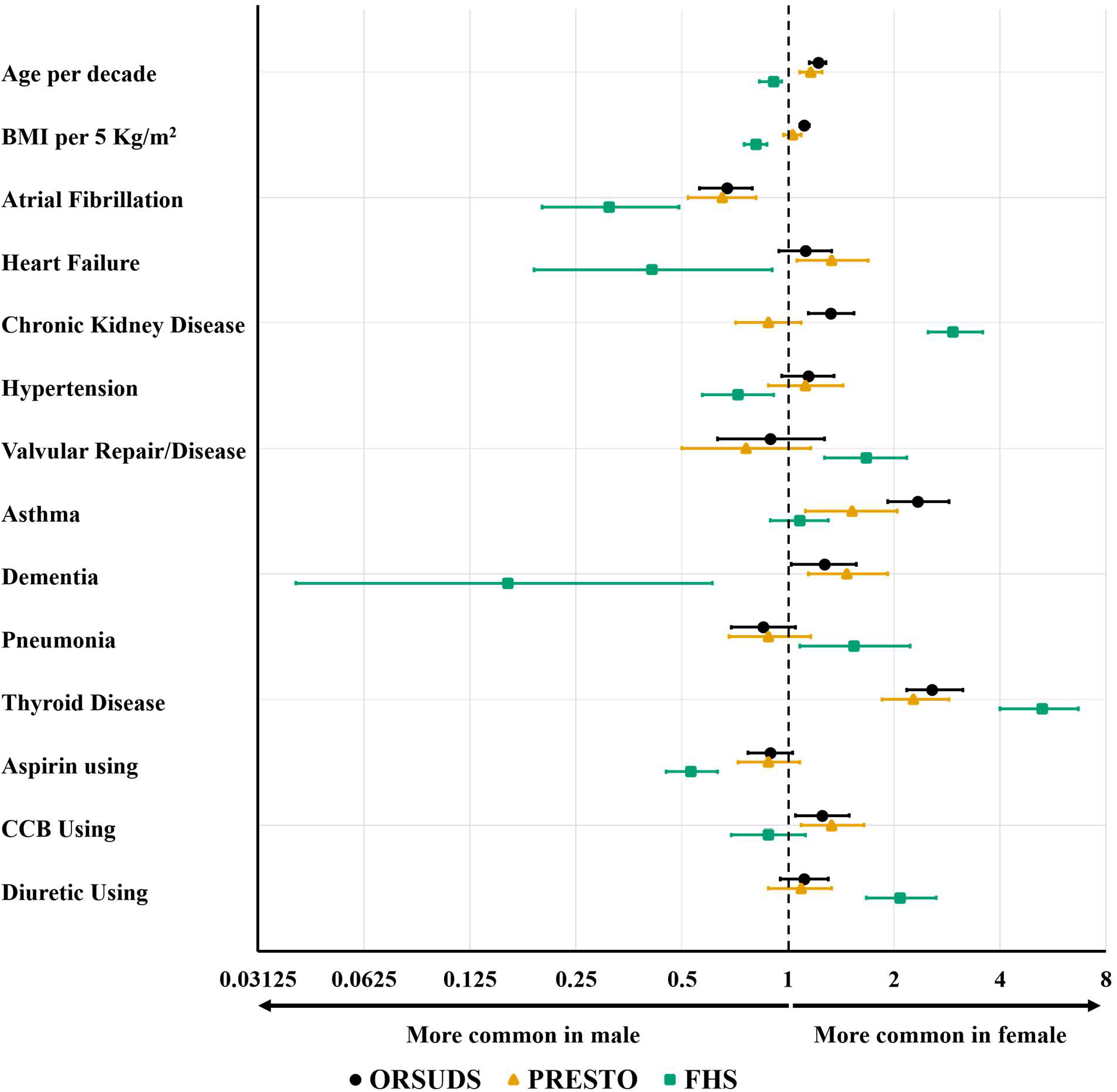
Heterogeneity in Sex Differences of Clinical Characteristics Between the Discovery Cohort (ORSUDS) and the Community-Based Reference Cohort, (FHS) This figure presents the 14 of 25 clinical and medication features that showed significant heterogeneity in associations with sex between the discovery cohort (ORSUDS) and the community-based reference cohort (FHS), based on false discovery rate (FDR)–adjusted P values from Cochran’s tests. PRESTO results are shown for descriptive comparison with ORSUDS. For eight features—age, body mass index (BMI; kg/m²), heart failure, hypertension, valvular disease or repair, dementia, pneumonia, and calcium channel blocker (CCB) use—the estimated associations with sex were in opposite directions in ORSUDS and FHS. For the remaining six features—atrial fibrillation, chronic kidney disease, asthma, thyroid disease, aspirin use, and diuretic use—significant heterogeneity reflected differences in the magnitude of the associations, with consistent directions across the two cohorts. The other 11 features showed no statistically significant heterogeneity between ORSUDS and FHS.

Significant ORSUDS–FHS heterogeneity was present for 5 of the 10 ECG features (Figure 3). The most pronounced direction reversal involved prolonged QTc, which was strongly male- associated in ORSUDS but female-associated in FHS (OR 0.45 versus 1.64; Q=31.41, FDR- adjusted p < 0.001). ECG-defined LVH was strongly female-associated in ORSUDS but showed a nonsignificant male-directed estimate in FHS (OR 3.03 versus 0.83; Q=41.62, FDR-adjusted p < 0.001). Heterogeneity in intraventricular conduction delay, delayed intrinsic deflection, and abnormal QRS axis reflected substantially stronger male associations in FHS. In contrast, the sex associations for bradycardia, prolonged PR interval, premature contractions, tachycardia, and atrial enlargement showed no significant between-cohort heterogeneity (Table S7).

**Figure 3.**
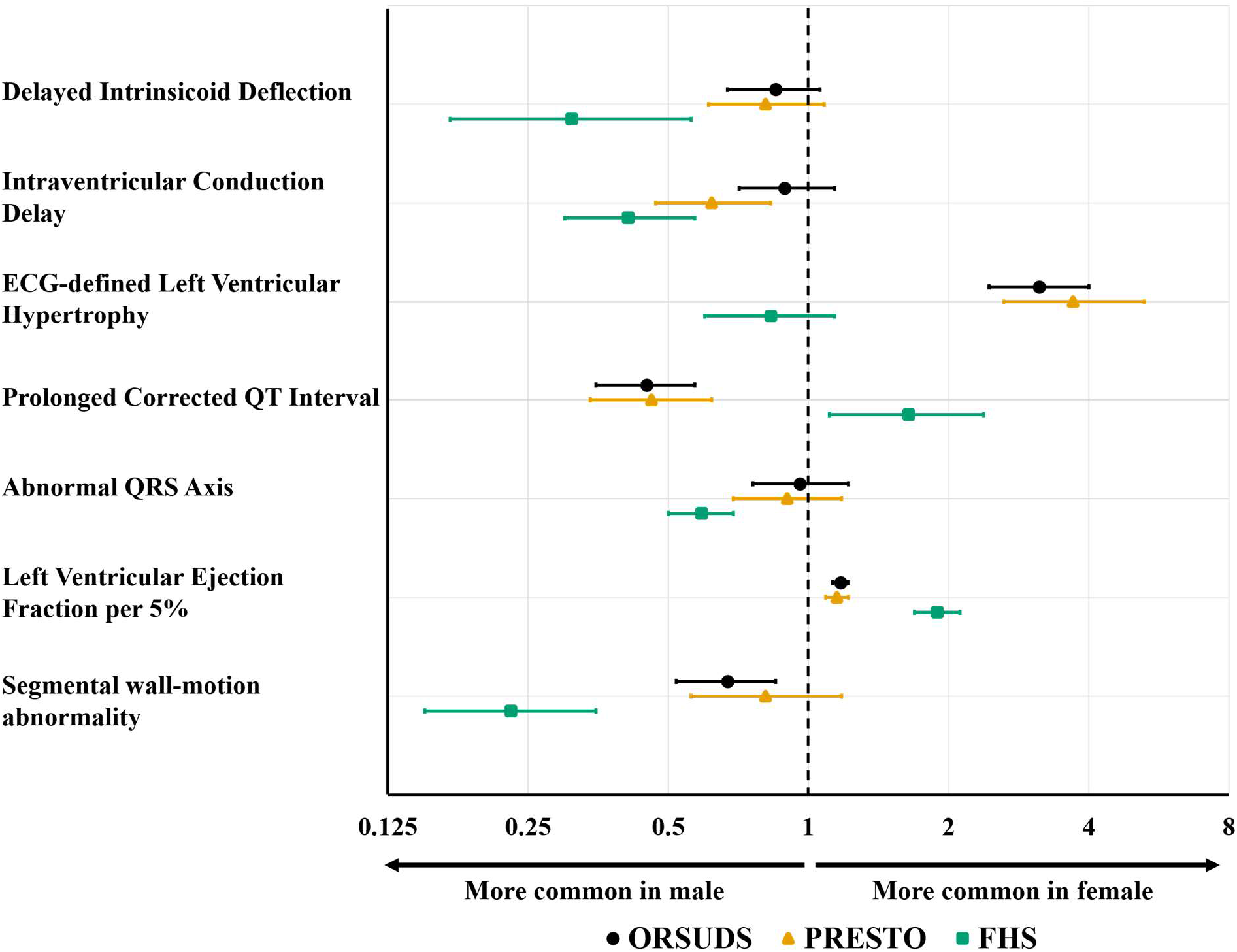
Heterogeneity in Sex Differences of ECG and Echo features Between the Discovery Cohort (ORSUDS) and the Community-Based Reference Cohort, (FHS) This figure presents associations with sex for electrocardiographic (ECG) and echocardiographic features in the discovery cohort (ORSUDS) and the community-based reference cohort (FHS). Heterogeneity was assessed using Cochran’s tests with false discovery rate (FDR)–adjusted P values. PRESTO results are shown for descriptive comparison with ORSUDS. Significant heterogeneity was observed for 5 of 10 ECG features. Prolonged corrected QT interval (QTc) showed the most pronounced directional reversal, with an association with male sex in ORSUDS and female sex in FHS. ECG-defined left ventricular hypertrophy (LVH) was associated with female sex in ORSUDS, whereas the FHS estimate was nonsignificant male-directed. Heterogeneity in intraventricular conduction delay, delayed intrinsic deflection, and abnormal QRS axis reflected stronger male associations in FHS. Regarding the echocardiography subset, higher left ventricular ejection fraction (LVEF) was associated with female sex in both cohorts, but the association was stronger in FHS. Segmental wall-motion abnormalities were associated with male sex in both cohorts, also more strongly in FHS

Significant ORSUDS–FHS heterogeneity was observed for 2 of the 7 echocardiographic features: LVEF and segmental wall-motion abnormalities (Figure 3). The association between higher LVEF and female sex was present in both cohorts but was substantially stronger in FHS (OR per 5% increase 1.17 in ORSUDS versus 1.89 in FHS; Q=61.88, FDR-adjusted p < 0.001). Similarly, segmental wall-motion abnormalities were male-associated in both cohorts but more strongly associated with male sex in FHS (OR 0.70 versus 0.23; Q=18.42, FDR-adjusted p < 0.001). There was no evidence of significant heterogeneity for echocardiographic LVH, left atrial enlargement, LV enlargement, mitral valve abnormalities, or aortic valve abnormalities (Table S7).

## Discussion

In this community-based study, we identified distinct pre-arrest profiles among women and men with SCA across three complementary domains: clinical profile, ECG features, and echocardiographic findings. SCA in women was more strongly associated with non-cardiac comorbidities, LVH, preserved LVEF, and mitral valve abnormalities. In contrast, SCA in men was characterized by a greater burden of cardiac disease, behavioral risk factors, major ECG abnormalities, and left ventricular systolic dysfunction. These patterns were externally validated in PRESTO, with 95.1% of tested associations showing no significant FDR-adjusted between- cohort heterogeneity, supporting their reproducibility across geographically and demographically distinct SCA populations. In contrast, approximately half of the sex-associated differences observed among SCA cases diverged from corresponding sex patterns in the community cohort, the Framingham Heart Study: about one-quarter were directionally discordant, and another one- quarter differed primarily in magnitude. The observed heterogeneity between the SCA and community cohorts suggest that the observed sex-associated SCA profiles do not simply reflect background population differences but may instead represent potentially distinct pathways of SCD vulnerability in women versus men.

Specifically, women with SCA were older and demonstrated a broader non-cardiac comorbidity profile, with higher burdens of endocrine, immune, respiratory, pain-related, and neuropsychiatric conditions. Although several of these conditions have an established female predominance in the general population, their association with female sex among SCA cases— particularly thyroid disease, autoimmune disease, asthma, chronic pain, dementia, and mood disorder—has not been comprehensively described and represents a novel contribution of this study. Some of these associations may partly reflect background population sex differences, whereas others may be more characteristic of the clinical profile preceding SCA. Female cases also had more frequent ECG-defined LVH. The LVH finding is also consistent with the Fingesture study, in which LVH was the only ECG abnormality more common among women with SCD.^12^ Our study extends this observation by demonstrating female enrichment of LVH on both ECG and echocardiography. Female cases were also more likely to have preserved LVEF and mitral valve abnormalities, consistent with prior SCA studies reporting less severe left ventricular systolic dysfunction among women.^26^ Collectively, these findings define a SCA for women with SCA characterized by non-cardiac comorbidities, hypertrophic remodeling, and preserved systolic function, which may be less readily recognized by conventional SCA profiles centered on documented CAD and reduced LVEF.

In contrast, men with SCA were characterized by a predominantly cardiovascular–arrhythmic phenotype across the clinical, electrical, and structural domains. Clinically, male cases had greater burdens of CAD, AF, peripheral vascular disease, and sleep apnea, together with greater exposure to behavioral risk factors, particularly heavy alcohol and illicit drug use. The predominance of CAD is consistent with previous studies, in which ischemic heart disease was more common among men than women with SCD.^3,4,6,12,27^ To our knowledge, although they are recognized risk factors for SCA, peripheral vascular disease and sleep apnea have not previously been emphasized as sex-associated features among SCA cases and therefore represent potentially novel components of the male clinical phenotype. The greater prevalence of sleep apnea in men with SCA despite significantly lower BMI than women with SCA could be partly explained by previous reports of more vulnerable anatomical and physiological characteristics (thick neck circumference and high neck-to-height ratio) relevant to sleep apnea along with established male predominance in the general population,^28,29^ whereas the enrichment of peripheral vascular disease may indicate broader vascular vulnerability relevant to SCA or may be influenced by under-recognition of vascular disease in women because of less typical clinical presentation.^30^ From the electrophysiological perspective, male cases were more likely to have prolonged PR, prolonged QTc, premature contractions, and bradycardia. The prolonged-QTc finding is consistent with Fingesture, which also reported higher prevalence of QTc prolongation and QRS abnormalities among men;^12^ our findings for prolonged PR and premature contractions extend the previously described electrical profile. Together, these electrical abnormalities are compatible with a greater burden of conduction-system disease, myocardial injury, and adverse remodeling among male cases. Echocardiographic findings further reinforced this pattern: male cases were more likely to have reduced LVEF and segmental wall-motion abnormalities. Collectively, these observations indicate that male SCA cases more often had recognized cardiovascular disease, electrical abnormalities, and systolic dysfunction before arrest—a pattern compatible with ischemic injury.

Comparison with FHS identified several sex-associated features whose patterns among SCA cases differed from those in the community cohort samples. The clearest divergence involved prevalence of prolonged QTc, which was male-enriched in both SCA cohorts but female- enriched in FHS. This finding is consistent with an early analysis reported from the ORSUDS study that evaluated determinants of QTc prolongation in a control group of individuals with coronary disease but no history of SCA.^31^ One possible explanation is that the greater burden of ischemic heart disease and left ventricular systolic dysfunction among male SCA cases may be accompanied by myocardial injury, adverse remodeling, and altered ventricular repolarization. ECG-defined LVH showed a complementary divergence: it was strongly female- associated in both SCA cohorts but tended to be male-associated in the FHS cohort. This finding is consistent with Fingesture, which reported a higher prevalence of ECG-LVH among women with SCD which was partly explained by the greater prevalence of primary myocardial fibrosis among women, raising the possibility that hypertrophic or fibrotic remodeling may contribute more prominently to the phenotype of women with SCA.^12^ Higher BMI also showed a directional reversal, being female-enriched in ORSUDS but male-enriched in FHS. One possible explanation is that male SCA cases more often exhibited ischemic heart disease and systolic dysfunction, which may be accompanied by disease-related weight loss and loss of lean mass, whereas female cases may be relatively enriched for obesity-related metabolic pathways. The latter is supported by a UK Biobank study in which the triglyceride–glucose–BMI index showed a stronger association with incident SCA in women than in men.^32^ This index reflects insulin resistance and metabolic burden, however, and does not establish a sex-specific effect of BMI itself. Hypertension and HF were also female-directed in both SCA cohorts but male-enriched in FHS. Together with the female enrichment of preserved LVEF and ECG defined LVH among SCA cases, this finding raises the possibility of a hypertrophic, preserved-EF phenotype among women which could be attributed to pressure overload. Moreover, several additional clinical, ECG, and echocardiographic features showed sex associations in the same direction in ORSUDS and FHS, but the magnitudes of these associations differed significantly between the two populations. For some features, the sex association was stronger among SCA cases, whereas for others it was stronger in FHS. These findings suggest that pathophysiologic mechanism associated with SCA may modify the strength of sex differences observed in the general population.

Collectively, our findings represent the first step in a three-stage strategy toward sex-specific SCA risk prediction. The first step is to establish whether women and men with SCA exhibit distinct pre-arrest profiles. The external reproducibility of these profiles and the divergence of selected features from corresponding community patterns provide evidence that clinically meaningful sex differences exist among SCA cases and identify candidate features for subsequent risk-factor studies. The second step is to determine whether these candidate features are associated with incident SCA differently in women and men. Previous women-focused studies have identified hypertension, diabetes, smoking, adiposity, prior MI, HF, LVH, and QT-related measures as predictors of SCA.^13–15^ However, comprehensive direct comparisons of the magnitude of these associations between women and men within the same source population remain limited. Such studies should compare women with and without SCA and men with and without SCA and formally test sex-by-feature interactions to determine whether each condition has a similar, stronger, weaker, or potentially opposite association with SCA by sex. The third and final step is to incorporate the resulting sex-specific risk factors into separate prediction models for women and men. Unlike a single model developed in a combined-sex population,^33–35^ separate models could use different predictors, predictor weights, thresholds, interactions, and nonlinear relationships that reflect the distinct profiles identified in the present study. The long-term objective is not simply to add sex as another variable to an existing SCA risk score, but to determine whether women and men require different predictors, different predictor weights, and ultimately different prediction models. The present study defines the sex-associated profiles needed to begin that process; subsequent studies must identify the corresponding sex-specific risk factors and translate them into externally validated risk scores.

Several limitations should also be considered. First, the primary case–case design in discovery cohort characterizes features associated with male versus female sex among individuals who experienced SCA; it cannot estimate sex-specific incident SCA risk, establish causal relationships, or determine the predictive value of the identified features. Second, FHS is a predominantly White, New England-based cohort, with most participants having a high school or greater education, which may limit the generalizability to the general population. Differences between FHS and ORSUDS participant selection and data-collection procedures may also have contributed to the observed heterogeneity, though findings were largely similar. Third, some features were unavailable or not identically defined across ORSUDS and FHS, and the cohort-specific models consequently differed in their covariate composition. Therefore, significant ORSUDS–FHS heterogeneity may partly reflect differences in measurement, variable availability, or model specification in addition to true differences between SCA cases and the reference cohort. Fourth, our findings were observational and we cannot exclude residual confounding. Fifth, although we used multiple imputations to address missing data, we acknowledge that the missing covariates may not have been missing at random. Finally, for SCA cases pre-arrest ECG and echocardiographic data were available only in clinically selected subsets and were obtained at varied intervals before SCA. This may have introduced selection and temporal measurement bias and may limit the generalizability of those findings to the entire SCA population.

Despite those limitations, this study has several important strengths. To our knowledge, it is the first community-based investigation to comprehensively characterize pre-arrest sex-associated profiles across clinical, ECG, and echocardiographic domains, evaluate their reproducibility in a geographically and demographically distinct SCA cohort, and formally compare them with a community-based reference cohort. The use of physician-adjudicated SCA cases, multivariable-adjusted models, independent external replication in PRESTO, and FDR-adjusted Cochran Q tests strengthened the robustness of the analyses. In addition, ORSUDS and PRESTO included a combined 2699 women with SCA, providing substantial female representation relative to most previous SCA studies.

## Conclusion

Women and men with SCA exhibited distinct pre-arrest profiles across clinical, ECG, and echocardiographic domains which were broadly reproducible in two separate SCA cohorts, ORSUDS and PRESTO. In contrast, many specific associations differed from corresponding sex patterns in the community-based FHS cohort. By defining and externally validating these sex-associated phenotypes, this study lays the foundations for identifying sex-specific SCA risk factors and ultimately developing and validating sex-specific prediction models toward the ultimate goal of enhancing SCA risk stratification and prevention.

## Ethical statement

The institutional review boards of Cedars-Sinai Medical Center, Oregon Health & Science University, Boston University Medical Campus, and all relevant hospitals/health systems have approved the study protocol.

## Acknowledgments

The authors would like to express their gratitude to the significant contribution of Portland, OR and Ventura, CA area residents, Global Medical Response, Portland/Gresham fire departments, and local area hospital systems (Oregon Health and Science University, Kaiser Permanente, Legacy Health, Adventist Health, Providence Health, Ventura County Medical Center, Los Robles Regional Medical Center, Community Memorial Hospital, Simi Valley Hospital, Dignity Health Care) and participants and the participants of the FHS (Framingham Heart Study) for their valuable contributions.

## Data availability statement

The data in this manuscript are from an ongoing study, and there is currently no IRB-approved mechanism by which this data will be deposited in a public repository. All analytical methods are included in this published article. De-identified participant data from ORSUDS and PRESTO will be made available after publication upon reasonable request to the corresponding author, following approval of a proposal and a signed data use agreement. FHS data cannot be redistributed but can be requested via the FHS Research Center, BioLINCC, or BioData Catalyst.

## Supplemental Material

Supplemental Methods

Supplemental Results

Tables S1–S7

References 1-3

## Abbreviations

SCA: Sudden Cardiac Arrest
CAD: Coronary Artery Disease
MI: Myocardial Infarction
HF: Heart Failure
LVH: Left Ventricular Hypertrophy
ORSUDS: Oregon Sudden Unexpected Death Study
PRESTO: Prediction of Sudden Death in Multi-Ethnic Communities
FHS: Framingham Heart Study
EMS: Emergency Medical Services
SD: Standard Deviation
CI: Confidence Interval
FDR: False Discovery Rate
BMI: Body Mass Index
CKD: Chronic Kidney Disease
AF: Atrial Fibrillation
LVEF: Left Ventricular Ejection Fraction

## References

1. Bogle BM, Ning H, Mehrotra S, Goldberger JJ, Lloyd-Jones DM. Lifetime Risk for Sudden Cardiac Death in the Community. J Am Heart Assoc. 2016;5. doi: 10.1161/jaha.115.002398

2. Niemeijer MN, van den Berg ME, Leening MJ, Hofman A, Franco OH, Deckers JW, Heeringa J, Rijnbeek PR, Stricker BH, Eijgelsheim M. Declining incidence of sudden cardiac death from 1990-2010 in a general middle-aged and elderly population: The Rotterdam Study. Heart Rhythm. 2015;12:123–129. doi: 10.1016/j.hrthm.2014.09.054

3. Skjelbred T, Rajan D, Svane J, Lynge TH, Tfelt-Hansen J. Sex differences in sudden cardiac death in a nationwide study of 54 028 deaths. Heart. 2022;108:1012–1018. doi: 10.1136/heartjnl-2021-320300

4. Butters A, Arnott C, Sweeting J, Winkel BG, Semsarian C, Ingles J. Sex Disparities in Sudden Cardiac Death. Circulation: Arrhythmia and Electrophysiology. 2021;14:e009834. doi: 10.1161/CIRCEP.121.009834

5. Reinier K, Dizon B, Chugh H, Bhanji Z, Seifer M, Sargsyan A, Uy-Evanado A, Norby FL, Nakamura K, Hadduck K, et al. Warning symptoms associated with imminent sudden cardiac arrest: a population-based case-control study with external validation. Lancet Digit Health. 2023;5:e763–e773. doi: 10.1016/s2589-7500(23)00147-4

6. Chugh SS, Uy-Evanado A, Teodorescu C, Reinier K, Mariani R, Gunson K, Jui J. Women have a lower prevalence of structural heart disease as a precursor to sudden cardiac arrest: The Ore-SUDS (Oregon Sudden Unexpected Death Study). J Am Coll Cardiol. 2009;54:2006–2011. doi: 10.1016/j.jacc.2009.07.038

7. Kotini-Shah P, Del Rios M, Khosla S, Pugach O, Vellano K, McNally B, Vanden Hoek T, Chan PS. Sex differences in outcomes for out-of-hospital cardiac arrest in the United States. Resuscitation. 2021;163:6–13. doi: 10.1016/j.resuscitation.2021.03.020

8. Blom MT, Oving I, Berdowski J, van Valkengoed IGM, Bardai A, Tan HL. Women have lower chances than men to be resuscitated and survive out-of-hospital cardiac arrest. European Heart Journal. 2019;40:3824–3834. doi: 10.1093/eurheartj/ehz297

9. Perman SM, Shelton SK, Knoepke C, Rappaport K, Matlock DD, Adelgais K, Havranek EP, Daugherty SL. Public Perceptions on Why Women Receive Less Bystander Cardiopulmonary Resuscitation Than Men in Out-of-Hospital Cardiac Arrest. Circulation. 2019;139:1060–1068. doi: 10.1161/CIRCULATIONAHA.118.037692

10. Reinier K, Stecker EC, Uy-Evanado A, Chugh HS, Binz A, Nakamura K, Sargsyan A, Jui J, Chugh SS. Sudden Cardiac Death as First Manifestation of Heart Disease in Women. Circulation. 2020;141:606–608. doi: 10.1161/CIRCULATIONAHA.119.044169

11. Tseng ZH, Ramakrishna S, Salazar JW, Vittinghoff E, Olgin JE, Moffatt E. Sex and Racial Differences in Autopsy-Defined Causes of Presumed Sudden Cardiac Death. Circ Arrhythm Electrophysiol. 2021;14:e009393. doi: 10.1161/circep.120.009393

12. Haukilahti MAE, Holmström L, Vähätalo J, Kenttä T, Tikkanen J, Pakanen L, Kortelainen ML, Perkiömäki J, Huikuri H, Myerburg RJ, Junttila MJ. Sudden Cardiac Death in Women. Circulation. 2019;139:1012–1021. doi: 10.1161/circulationaha.118.037702

13. Bertoia ML, Allison MA, Manson JE, Freiberg MS, Kuller LH, Solomon AJ, Limacher MC, Johnson KC, Curb JD, Wassertheil-Smoller S, Eaton CB. Risk factors for sudden cardiac death in post-menopausal women. J Am Coll Cardiol. 2012;60:2674–2682. doi: 10.1016/j.jacc.2012.09.031

14. Albert CM, Chae CU, Grodstein F, Rose LM, Rexrode KM, Ruskin JN, Stampfer MJ, Manson JE. Prospective study of sudden cardiac death among women in the United States. Circulation. 2003;107:2096–2101. doi: 10.1161/01.Cir.0000065223.21530.11

15. Mehta PK, Johnson BD, Kenkre TS, Eteiba W, Sharaf B, Pepine CJ, Reis SE, Rogers WJ, Kelsey SF, Thompson DV, et al. Sudden Cardiac Death in Women With Suspected Ischemic Heart Disease, Preserved Ejection Fraction, and No Obstructive Coronary Artery Disease: A Report From the Women’s Ischemia Syndrome Evaluation Study. J Am Heart Assoc. 2017;6. doi: 10.1161/jaha.117.005501

16. Reinier K, Chugh H, Kadiyala V, Sargsyan A, Uy-Evanado A, Nakamura K, Heckard E, Mathias M, Grogan T, Elashoff D, et al. Clinical Triage of Individuals With Warning Symptoms of Imminent Cardiac Arrest. Circ Arrhythm Electrophysiol. 2026;19:e014647. doi: 10.1161/CIRCEP.125.014647

17. Chugh SS, Jui J, Gunson K, Stecker EC, John BT, Thompson B, Ilias N, Vickers C, Dogra V, Daya M, et al. Current burden of sudden cardiac death: multiple source surveillance versus retrospective death certificate-based review in a large U.S. community. J Am Coll Cardiol. 2004;44:1268–1275. doi: 10.1016/j.jacc.2004.06.029

18. Fishman GI, Chugh SS, Dimarco JP, Albert CM, Anderson ME, Bonow RO, Buxton AE, Chen PS, Estes M, Jouven X, et al. Sudden cardiac death prediction and prevention: report from a National Heart, Lung, and Blood Institute and Heart Rhythm Society Workshop. Circulation. 2010;122:2335–2348. doi: 10.1161/circulationaha.110.976092

19. Dawber TR, Kannel WB, Lyell LP. An approach to longitudinal studies in a community: the Framingham Study. Ann N Y Acad Sci. 1963;107:539–556. doi: 10.1111/j.1749-6632.1963.tb13299.x

20. Dawber TR, Meadors GF, Moore FE, Jr. Epidemiological approaches to heart disease: the Framingham Study. Am J Public Health Nations Health. 1951;41:279–281. doi: 10.2105/ajph.41.3.279

21. Pope MK, Chugh H, Truyen T, Mathias M, Uy-Evanado A, Lin H, Atar D, Bosson N, Reinier K, Benjamin EJ, Chugh SS. Dynamic Risk Trajectories for Sudden Cardiac Arrest: The Role of Recurrent Cardiovascular Events. J Am Heart Assoc. 2026;15:e049853. doi: 10.1161/JAHA.125.049853

22. Kannel WB, Feinleib M, McNamara PM, Garrison RJ, Castelli WP. An investigation of coronary heart disease in families. The Framingham offspring study. Am J Epidemiol. 1979;110:281–290. doi: 10.1093/oxfordjournals.aje.a112813

23. Mongraw-Chaffin ML, Peters SAE, Huxley RR, Woodward M. The sex-specific association between BMI and coronary heart disease: a systematic review and meta-analysis of 95 cohorts with 1·2 million participants. Lancet Diabetes Endocrinol. 2015;3:437–449. doi: 10.1016/s2213-8587(15)00086-8

24. Woodward M. Rationale and tutorial for analysing and reporting sex differences in cardiovascular associations. Heart. 2019;105:1701–1708. doi: 10.1136/heartjnl-2019-315299

25. Benjamini Y, Hochberg Y. Controlling the False Discovery Rate: A Practical and Powerful Approach to Multiple Testing. Journal of the Royal Statistical Society: Series B (Methodological). 1995;57:289–300. doi: 10.1111/j.2517-6161.1995.tb02031.x

26. Stecker Eric C, Vickers C, Waltz J, Socoteanu C, John Benjamin T, Mariani R, McAnulty John H, Gunson K, Jui J, Chugh Sumeet S. Population-Based Analysis of Sudden Cardiac Death With and Without Left Ventricular Systolic Dysfunction. JACC. 2006;47:1161–1166. doi: 10.1016/j.jacc.2005.11.045

27. Albert CM, McGovern BA, Newell JB, Ruskin JN. Sex differences in cardiac arrest survivors. Circulation. 1996;93:1170–1176. doi: 10.1161/01.cir.93.6.1170

28. Won CHJ, Reid M, Sofer T, Azarbarzin A, Purcell S, White D, Wellman A, Sands S, Redline S. Sex differences in obstructive sleep apnea phenotypes, the multi-ethnic study of atherosclerosis. Sleep. 2020;43. doi: 10.1093/sleep/zsz274

29. Subramanian S, Jayaraman G, Majid H, Aguilar R, Surani S. Influence of gender and anthropometric measures on severity of obstructive sleep apnea. Sleep and Breathing. 2012;16:1091–1095. doi: 10.1007/s11325-011-0607-9

30. Pabon M, Cheng S, Altin SE, Sethi SS, Nelson MD, Moreau KL, Hamburg N, Hess CN. Sex Differences in Peripheral Artery Disease. Circ Res. 2022;130:496–511. doi: 10.1161/circresaha.121.320702

31. Chugh SS, Reinier K, Singh T, Uy-Evanado A, Socoteanu C, Peters D, Mariani R, Gunson K, Jui J. Determinants of prolonged QT interval and their contribution to sudden death risk in coronary artery disease: the Oregon Sudden Unexpected Death Study. Circulation. 2009;119:663–670. doi: 10.1161/CIRCULATIONAHA.108.797035

32. Zhang S, Liu W, Xu B, Wang S, Du Z, Cheng W. Association of triglyceride glucose index and triglyceride glucose-body mass index with sudden cardiac arrest in the general population. Cardiovascular Diabetology. 2024;23:173. doi: 10.1186/s12933-024-02275-2

33. Chugh SS, Reinier K, Uy-Evanado A, Chugh HS, Elashoff D, Young C, Salvucci A, Jui J. Prediction of Sudden Cardiac Death Manifesting With Documented Ventricular Fibrillation or Pulseless Ventricular Tachycardia. JACC: Clinical Electrophysiology. 2022;8:411–423. doi: 10.1016/j.jacep.2022.02.004

34. Truyen TTTT, Lin H, Mathias M, Chugh H, Reinier K, Benjamin EJ, Chugh SS. Validation of a Novel Risk Prediction Score for Sudden Cardiac Death in the Framingham Heart Study. Circulation: Arrhythmia and Electrophysiology. 2025;18:e013647. doi: 10.1161/CIRCEP.124.013647

35. Bogle BM, Ning H, Goldberger JJ, Mehrotra S, Lloyd-Jones DM. A Simple Community-Based Risk-Prediction Score for Sudden Cardiac Death. Am J Med. 2018;131:532–539.e535. doi: 10.1016/j.amjmed.2017.12.002

